# Epidemiology of Respiratory Viruses in Acute Respiratory Illnesses in Malaysia: Patterns, Seasonality and Age Distribution

**DOI:** 10.1101/2022.03.03.22271672

**Authors:** Yoke Lee Low, Shin Yee Wong, Hor Eric Kim Lee, Mohd. Hareeff Muhammed

## Abstract

**Objectives:** Acute Respiratory Infections (ARIs) are one of the leading causes of childhood morbidity and mortality worldwide. However, there is limited surveillance data on the epidemiological burden of respiratory pathogens in tropical countries like Malaysia. This study aims to estimate the prevalence of respiratory pathogens causing ARIs among children aged <18 years old in Malaysia and their epidemiological characteristics.

**Methods:** Nasopharyngeal swab specimens received at 12 laboratories located in different states of Malaysia from 2015-2019 were studied. Detection of 18 respiratory pathogens were performed using multiplex PCR.

**Results:** Data from a total of 23,306 paediatric patients who presented with ARI over a five-year period was studied. Of these, 18538 (79.5%) were tested positive. The most prevalent respiratory pathogens detected in this study were enterovirus/ rhinovirus (6837/ 23000; 29.7%), influenza virus (5176/ 23000; 22.5%) and respiratory syncytial virus (RSV) (3652/ 23000; 15.9%). Throughout the study period, RSV demonstrated the most pronounce seasonality; peak infection occurred during July to September. Whereas the influenza virus was detected year-round in Malaysia. No seasonal variation was noted in other respiratory pathogens. The risk of RSV hospitalisation was found to be significantly higher in children aged less than two years old, whereas hospitalisation rates for the influenza virus peaked at children aged between 3-6 years old.

**Conclusions:** This study provides insight into the epidemiology and the seasonality of the causative pathogens of ARI among the paediatric population in Malaysia. Knowledge of seasonal respiratory pathogens epidemiological dynamics will facilitate the identification of a target window for vaccination.

## Introduction

Acute respiratory infections (ARIs) are one of the leading causes of childhood morbidity and mortality worldwide. It is estimated that each year more than 2 million children die from ARIs with 70% of the mortality occurring in Africa and Southeast Asia [1]. Most ARIs are of viral origin in children less than 5 years old [2]. The association between ARIs and different viral pathogens ranges between 40% to 90% [3-6]. Influenza A and B virus, enterovirus/ rhinovirus, respiratory syncytial virus (RSV), adenovirus and parainfluenza virus are among the common pathogens detected [7-8]. With more and more new pathogens like human metapneumovirus (hMPV) and human bocavirus reported over the decades [9], there is an increased urgency to study the epidemiology of respiratory tract pathogens in order to facilitate the planning of appropriate disease prevention and control strategies.

Detection of the specific respiratory viral pathogens in ARIs is essential for the proper management of patients and to avoid the unnecessary use of antibiotics [10]. Over the past decades, culture sensitivity, and serology testing has been the mainstay of clinical laboratory diagnosis for respiratory pathogen detection. Increasing use of molecular techniques including polymerase chain reaction (PCR) has improved the sensitivity of detecting and identifying respiratory pathogens, providing new information on the epidemiology of ARIs besides contributing to a better understanding of the seasonality of the causative pathogens and their association with certain clinical manifestations [11-12].

Respiratory syncytial virus (RSV) and influenza virus are recognised as the most significant causes of ARIs in children worldwide [13]. Nearly all children are infected with RSV at least once during the first two years of life, with approximately two-third being infected by the end of their first year. The severity of RSV infections decreases with age, with a mild-to-moderate upper respiratory tract infection in healthy adults. [14]. Unlike RSV, influenza can affect people in any age group. Influenza complications and hospitalisation however are more common in childhood, especially in children age < 5 years, adults aged ≥ 65 years and those with chronic disease [15-16]. In most temperate climates, influenza and RSV activity consistently peaked during winter months while the timing of epidemics was found to be more diverse in the tropics [17].

Despite the burden of ARIs on morbidity and mortality in children, only limited data is available from countries in tropical and subtropical areas [18-20]. In order to determine the ongoing burden of viral-ARIs in the Malaysian paediatric population, a more representative surveillance data is needed. This will help in the implementation of prevention and treatment strategies for the management of ARIs in children. Thus, our study is aimed at evaluating the epidemiology of respiratory pathogens causing ARIs among children aged < 18 years in Malaysia, a tropical country north of equator. We further characterise the seasonal pattern and age-specific differences in influenza and RSV activity.

## Method

### Subject Enrolment and Sampling

This retrospective database review study was conducted in Pantai Premier Pathology, Kuala Lumpur. Nasopharyngeal/ nasal swab specimens received at 12 different laboratories located in different states of Malaysia from January 2015 to December 2019 were analysed. Swab samples were collected from paediatric in- and out-patients (age ≤18) who presented with signs and symptoms of respiratory tract infections including runny or stuffy nose, sore throat, cough, chills, fatigue and muscle ache. Nasopharyngeal specimens were obtained from these patients according to standard technique and placed in viral transport media. The requests for multiplex PCR testing were at the discretion of the physicians.

### Ethical Statement

Ethic approval for this study was obtained from Pantai Hospital Kuala Lumpur Research and Ethics Committee (Ethics Approval Number: PHKL-EC-2019-0012).

### Multiplex Polymerase Chain Reaction

Nasopharyngeal/ nasal samples were analysed by multiplex PCR method using Luminex NxTAG respiratory pathogen panel (Luminex Molecular Diagnostics, Toronto), BioFire FilmArray respiratory panel RP, RP2 (BioFire Diagnostics, Utah) or QIAstat-Dx respiratory panel V2 (Qiagen, Hilden) according to the manufacturer’s instructions. Pathogens excluded from the analysis due to inherent differences in the reagent kits used, include bocavirus, *Bordetella pertussis, Chlamydophila pneumoniae* and MERS coronavirus. 18 respiratory virus subtypes including non-specific influenza A, H1 subtype, H1N1(2009) subtype, H3 subtype, influenza B, parainfluenza1, parainfluenza2, parainfluenza3, parainfluenza4, coronavirus NL63, coronavirus HKU1, coronavirus 229E, coronavirus OC43, human metapneumovirus (hMPV), adenovirus, enterovirus/ rhinovirus, respiratory syncytial virus and one bacterial *Mycoplasma pneumoniae* were analysed. When more than one virus was detected simultaneously from the same patient, the findings will be recorded as multiple infections.

### Data Collection

Demographic data including medical record number, age and gender, respiratory sample results including the type of virus and the date of sample collection were retrieved from the laboratory database. Data were cleaned, screened from errors and duplication prior to data analysis.

### Statistical Analysis

Data was analysed with Statistical Package for Social Science (SPSS) version 26.0. Descriptive statistics (e.g frequency and percentage) were used for demographic characteristics. ANOVA and Kruskal-Wallis test is used to analyse non-normal distributed data. A *p*-value <0.05 was considered statistically significant.

## Results

### Demographic Characteristics of Study Population

During the five-year period from 2015–2019, specimens from a total of 23,306 paediatric patients who presented with ARI were analysed. Table 1 outlines the social-demographic variables of all the sample population. On average, 51.9% of the patients were male. The mean (±SD) age for the study population was 2.9 (±3.1) years with the majority of the patients (36.1%) in the age group of 1-2 years old. 5391 (23.1%) of the patients were less than one year old, 8418 (36.1%) were between 1-2 years old, 6763 (29.0%) were 3-6 years old and 2734 (11.7%) were more than 7 years old. The numbers of samples received at the laboratory have increased 5-fold in 5 years with the compounded annual growth rate (CAGR) of 50%, from 1680 in 2015 to 8520 in 2019.

**Table 1.** Demographic data of the paediatric population analysed from year 2015 to 2019

|  | 2015 | 2016 | 2017 | 2018 | 2019 | Total |
| --- | --- | --- | --- | --- | --- | --- |
| **Gender** |  |  |  |  |  |  |
| Male | 912 (54.3%) | 1402 (55.5%) | 2221 (55.6%) | 3650 (55.4%) | 3904 (45.8%) | 12089 (51.9%) |
| Female | 768 (45.7%) | 1124 (44.5%) | 1773 (44.4%) | 2936 (44.6%) | 4616 (54.2%) | 11217 (48.1%) |
| **Age** |  |  |  |  |  |  |
| <1 | 449 (26.7%) | 681 (27.0%) | 1043 (26.1%) | 1435 (21.8%) | 1783 (20.9%) | 5391 (23.1%) |
| 1 - 2 | 570 (33.9%) | 909 (36.0%) | 1512 (37.9%) | 2384 (36.2%) | 3043 (35.7%) | 8418 (36.1%) |
| 3 - 6 | 441 (26.3%) | 666 (26.4%) | 1078 (27.0%) | 2013 (30.6%) | 2565 (30.1%) | 6763 (29.0%) |
| 7 - 12 | 189 (11.3%) | 240 (9.5%) | 324 (8.1%) | 668 (10.1%) | 908 (10.7%) | 2329 (10.0%) |
| 13-17 | 31 (1.8%) | 30 (1.2%) | 37 (0.9%) | 86 (1.3%) | 221 (2.6%) | 405 (1.7%) |
| Total | 1680 (100.0%) | 2526 (100.0%) | 3994 (100.0%) | 6586 (100.0%) | 8520 (100.0%) | 23306 (100.0%) |

### Prevalence of Respiratory Pathogens

Of the 23,306 swab specimens received, 18538 (79.5%) were tested positive. Notably the total number of pathogens detected increased gradually from 1452 in year 2015 to 8965 in year 2019 (Table 2). A total of 23000 pathogens were detected by multiplex PCR. 5 predominant respiratory pathogens detected in this study were enterovirus/ rhinovirus (6837/ 23000; 29.7%), followed by influenza virus (5176/ 23000; 22.5%), RSV (3652/ 23000; 15.9%), adenovirus (2637/ 23000; 11.5%), and parainfluenza virus (2333/ 23000; 10.1%); Other less frequently detected respiratory pathogens including hMPV (819/ 23000; 3.6%) and *Mycoplasma pneumoniae* (338/ 23000; 1.5%).

**Table 2.** Prevalence of respiratory pathogens detected from year 2015 to 2019

|  | **2015** | **2016** | **2017** | **2018** | **2019** | **Total** |
| --- | --- | --- | --- | --- | --- | --- |
| Adenovirus | 234 (16.1%) | 190 (9.1%) | 479 (12.2%) | 918 (14.0%) | 816 (9.1%) | 2637 (11.5%) |
| Coronavirus | 39 (2.7%) | 73 (3.5%) | 182 (4.6%) | 195 (3.0%) | 330 (3.7%) | 819 (3.6%) |
| Human Metapneumovirus | 117 (8.1%) | 128 (6.1%) | 279 (7.1%) | 302 (4.6%) | 382 (4.3%) | 1208 (5.3%) |
| Enterovirus/ Rhinovirus | 465 (32.0%) | 579 (27.7%) | 1037 (26.4%) | 1968 (30.0%) | 2788 (31.1%) | 6837 (29.7%) |
| Influenza A | 178 (12.3%) | 166 (7.9%) | 608 (15.5%) | 833 (12.7%) | 1374 (15.3%) | 3159 (13.7%) |
| Influenza B | 38 (2.6%) | 250 (12.0%) | 269 (6.9%) | 620 (9.4%) | 840 (9.4%) | 2017 (8.8%) |
| Mycoplasma pneumoniae | 0 (0%) | 21 (1.0%) | 34 (0.9%) | 79 (1.2%) | 204 (2.3%) | 338 (1.5%) |
| Parainfluenza | 118 (8.1%) | 184 (8.8%) | 365 (9.3%) | 678 (10.3%) | 988 (11.0%) | 2333 (10.1%) |
| Respiratory Syncytial Virus | 263 (18.1%) | 500 (23.9%) | 672 (17.1%) | 974 (14.8%) | 1243 (13.9%) | 3652 (15.9%) |
| **Total** | 1452 (100.0%) | 2091 (100.0%) | 3925 (100.0%) | 6567 (100.0%) | 8965 (100.0%) | 23000 (100.0%) |

**Table 3.**
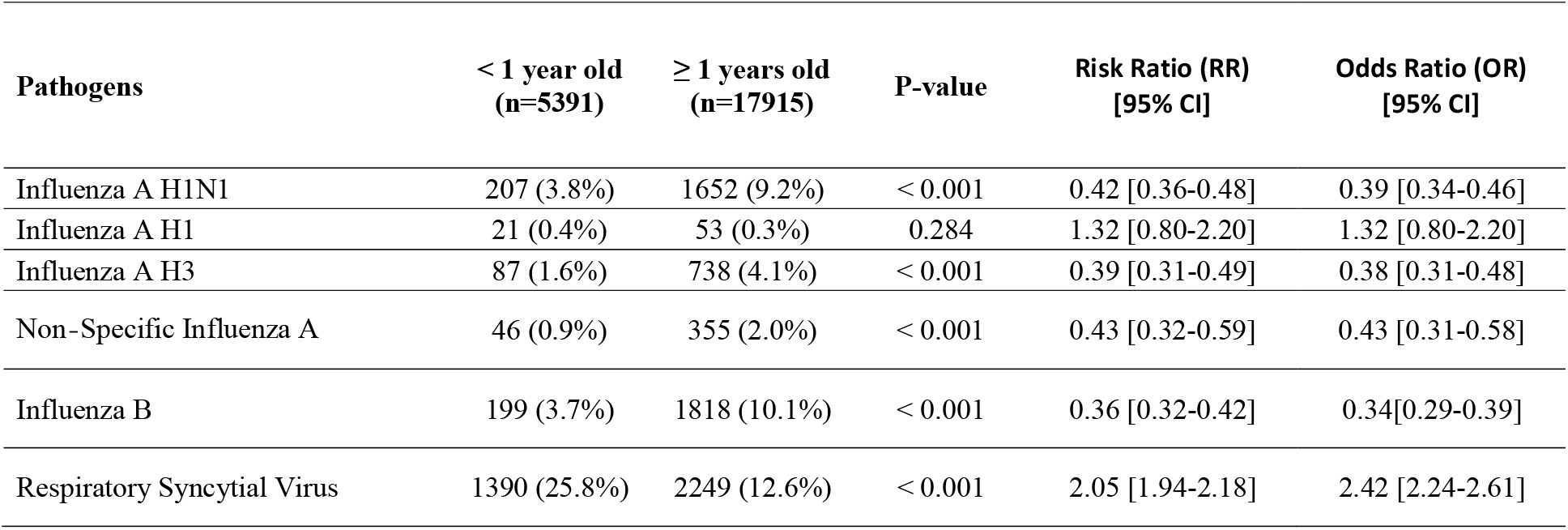
Distribution of Influenza A, Influenza B and Respiratory Syncytial Virus in different age group

### Percentage Positivity of Pathogens Detected in Different Paediatric Age Groups

Patients were divided into five different age groups namely, <1, 1-2, 3-6, 7-12 and 13-17 years old (Figure 1). Two respiratory pathogens found commonly in children below one year old (p-value <0.001) with the prevalence of 42.1% and 26.6% for enterovirus/ rhinovirus and RSV, respectively. On the other hand, influenza virus (peaked at 7 -12 years old, 53.3%), *Mycoplasma pneumoniae* (peaked at 13-17 years old, 7.2%) were more prevalent in older age groups (p-value <0.001). Prevalence of other respiratory pathogens peaked at different age groups: adenovirus (3-6 years old, 16.7%), coronavirus (<1 year old, 5.4%), hMPV (1-2 years old, 8.2%) and parainfluenza virus (<1 year old, 14.6%).

**Figure 1.**
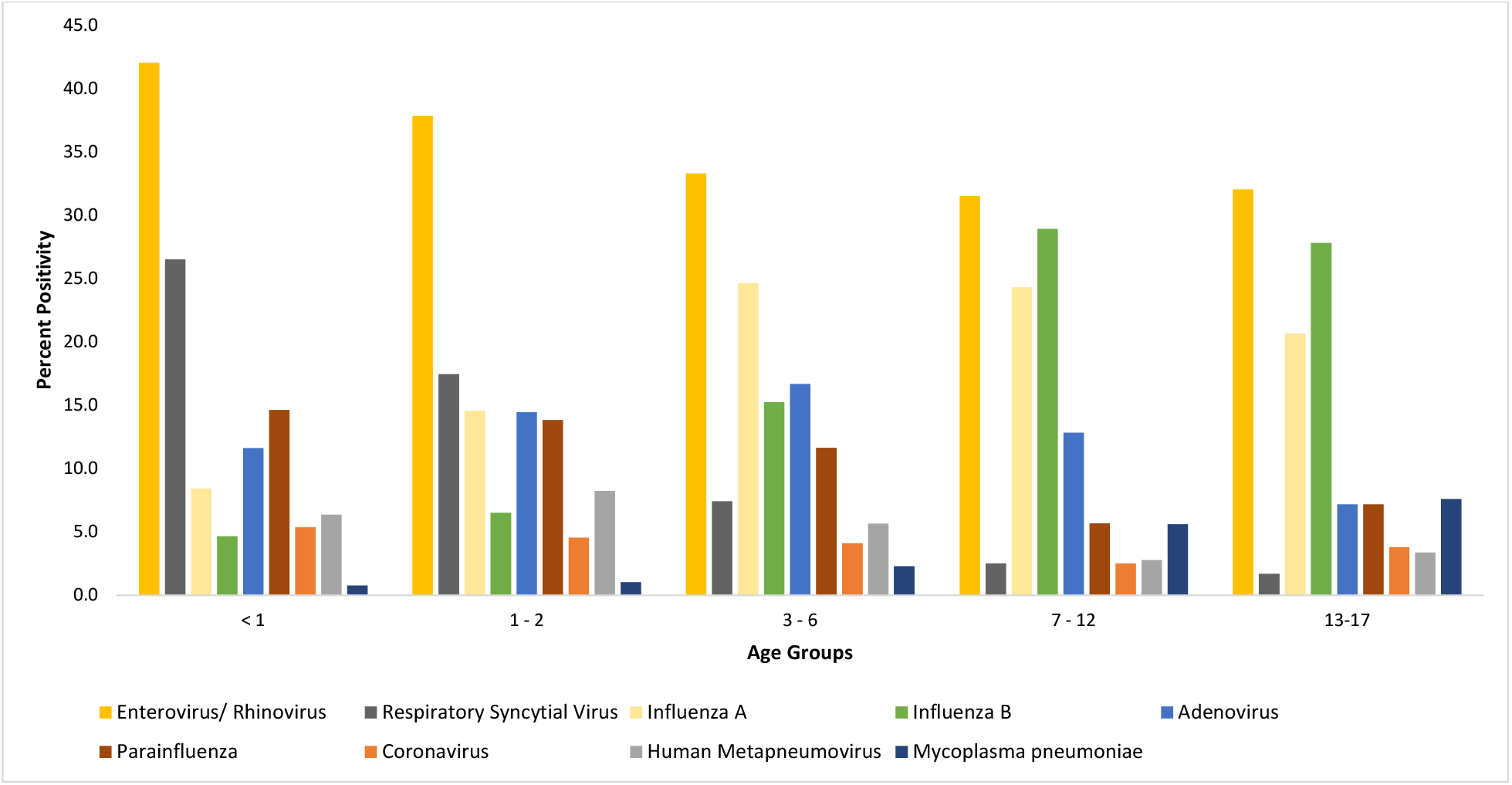
The percentage positivity of respiratory pathogens detected in different paediatric age groups

### Co-Infections Detected in the Samples

Co-infections were detected in 4242 samples (Figure 2), with a detection rate of 18.2% (4224/ 23306) of total specimens and 22.9% (4242/18538) of positive samples observed. Among the respiratory pathogens, enterovirus/ rhinovirus (76.8%), influenza B (72.1%) and RSV (69.5%) were predominantly detected as a single pathogen. Whereas coronavirus (59.6%) and Adenovirus (50.3%) were more commonly detected as co-infections. Co-infections were found to be most prevalent in children less than 1 years old (20.0%), while it was least common in elder children age 13-17 years old (7.2%). This difference was statistically significant (p-value <0.001)

**Figure 2.**
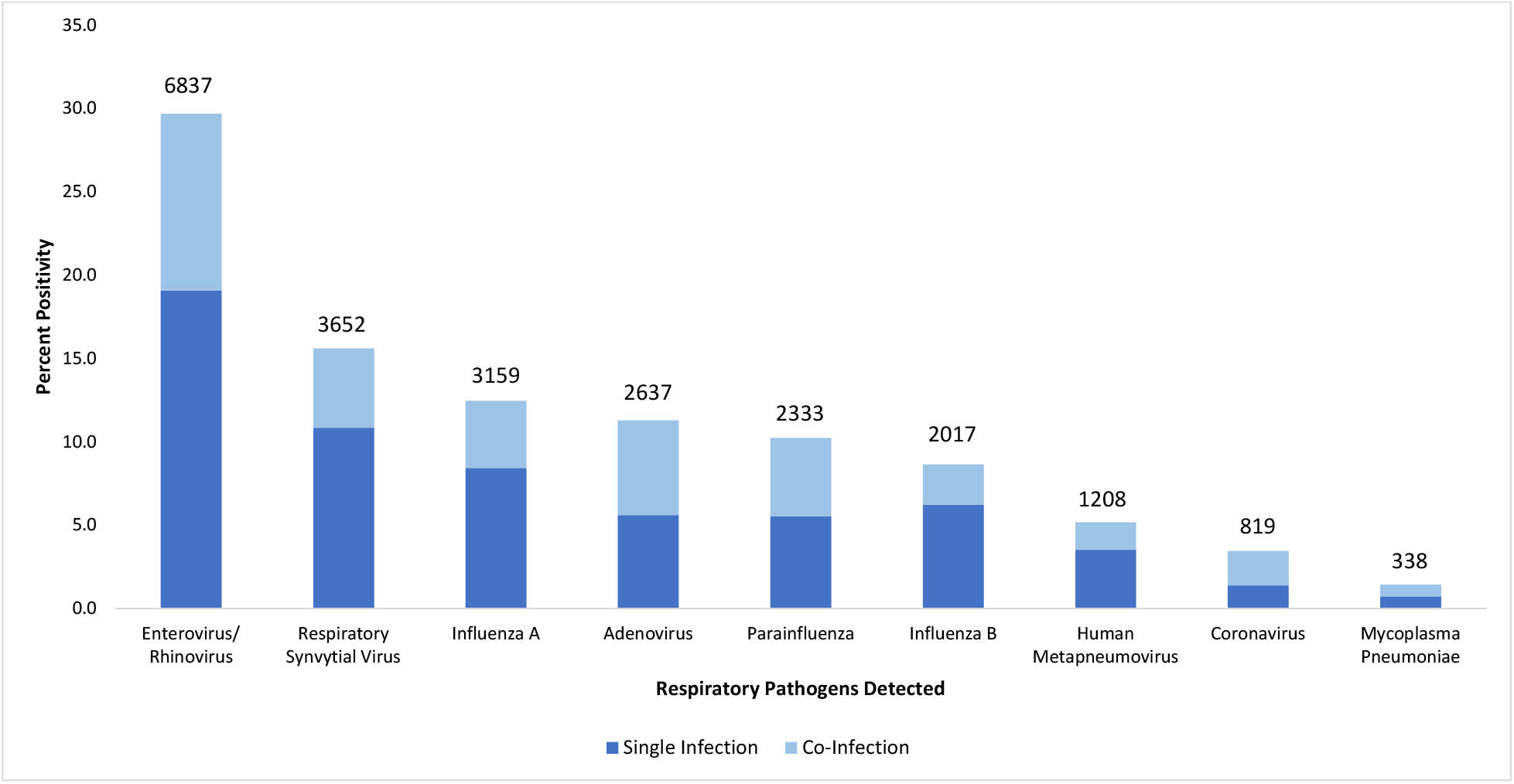
Percentage positivity of single and co-infections detected in each respiratory pathogen

### Seasonal Activity of Respiratory Pathogens

The seasonal distribution of the respiratory pathogens from January 2015 to December 2019 is shown in Figure 3. Overall positivity detection rate increased gradually year by year. Respiratory pathogens were detected throughout the year. Throughout the five-year study period, RSV demonstrated the most pronounced seasonality, with peak infection occurring from July to September (Figure 4). No seasonal variation was noted for influenza, enterovirus/ rhinovirus, adenovirus, coronavirus, parainfluenza virus, hMPV and *Mycoplasma Pneumoniae*. These viruses are detected year-round in Malaysia

**Figure 3.**
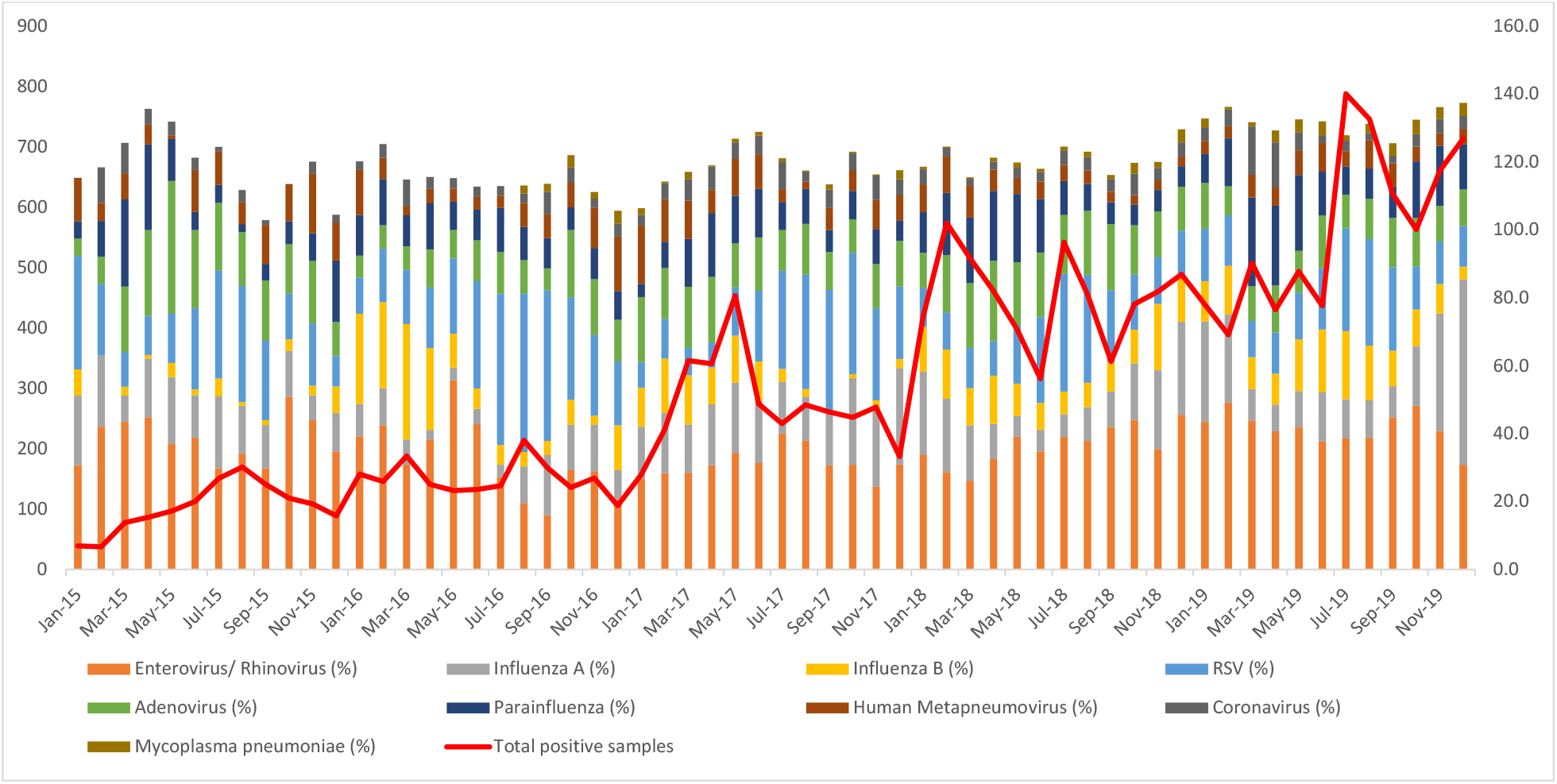
Monthly distribution of respiratory pathogens detected from January 2015 to December 2019

**Figure 4.**
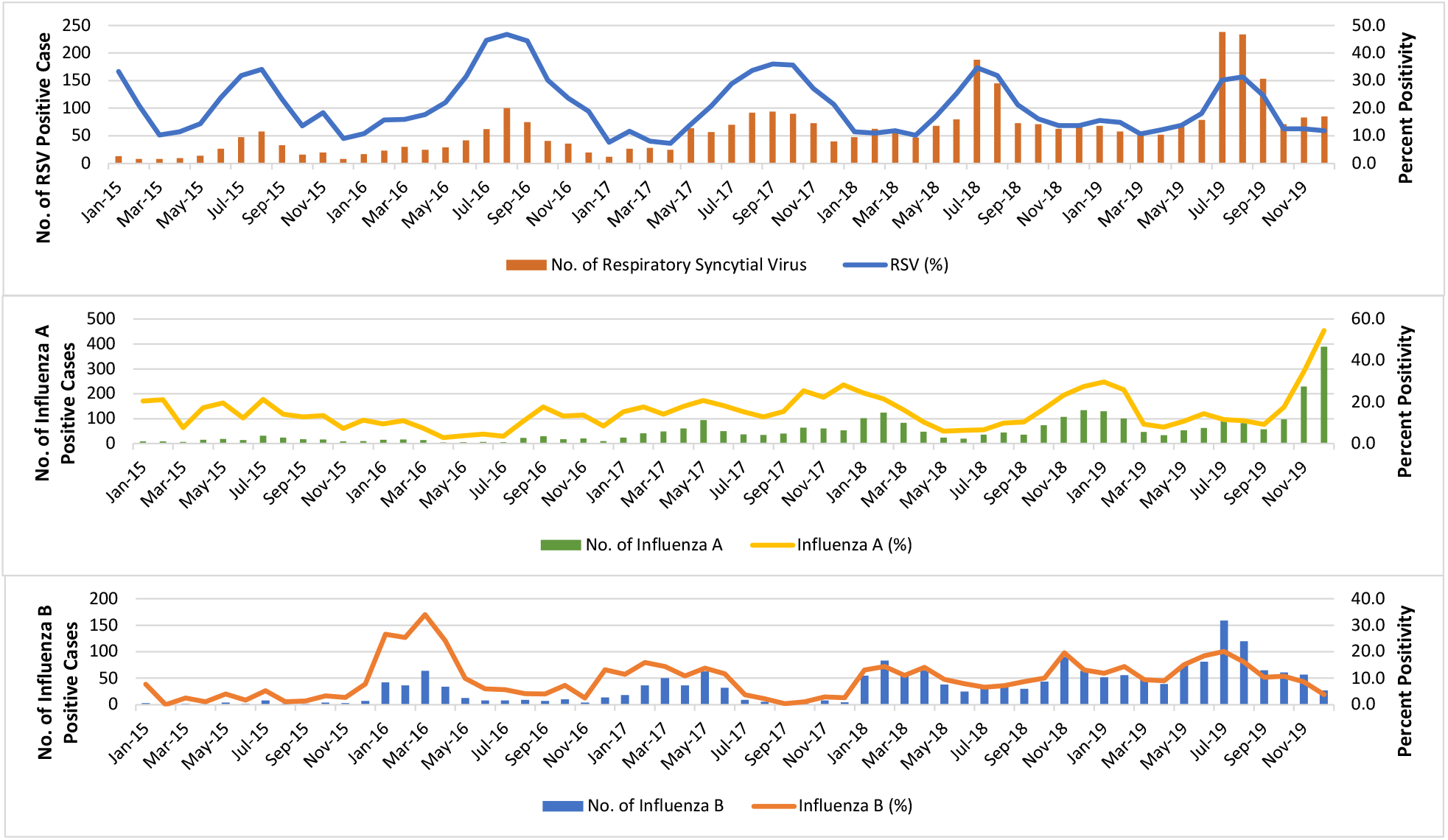
Monthly distribution of Influenza A, Influenza B and Respiratory Syncytial Virus from January 2015 to December 2019

### Age-Dependent Distribution of RSV and Influenza Infections

RSV (1390/5391) was predominantly detected in children less than 1-year old [P=<0.001, RR=2.05 (1.94-2.18), OR=2.42 (2.24-2.61)]. On the contrary, influenza A H1N1 [P=<0.001, RR=0.42 (0.36-0.48), OR=0.39 (0.34-0.46)], influenza A H3 [P=<0.001, RR=0.39 (0.31-0.49), OR=0.38 (0.31-0.48)], non-specific influenza A [P=<0.001, RR=0.43 (0.32-0.59), OR=0.43 (0.31-0.58)] and influenza B [P=<0.001, RR=0.36 (0.32-0.42), OR=0.34 (0.29-0.39)] were found to be more prevalent in children ≥1-year old.

### Age-Specific Hospitalisation Rates in Influenza and RSV

The risk of RSV hospitalisation was found to be significantly higher in children aged less than 2 years old; 42.3% in children less than 1 year old and 42.2% in children 1–2 years old respectively (Figure 5). RSV hospitalisation rates decreased with increasing age and did not contribute to any hospitalisations in children age group 13–17 years old. Influenza A and B share a similar pattern where hospitalisation rate was found to be more prevalent in children less than 6 years old, with the peak at 3-6 years old (influenza A=43.2%, influenza B=39.5%).

## Discussion

The Malaysian Ministry of Health reports that disease in the respiratory system is one of the major causes of hospitalisation in 2018, 13.86% and 18.73% in government and private hospitals, respectively [21]. However, there is limited epidemiological data as to the causative pathogens available in tropical countries, like Malaysia. This could be partly attributed to a lack of diagnostic capacity for viral detection. Previous studies using immunofluorescence (IF) and viral cultures had poor viral detection rates thus considerably underestimating the true burden of viral respiratory infections in Malaysia [22-23]. Real-time PCR offers a more advanced application, allowing the detection of a variety of viral and bacterial pathogens within hours. Rapid identification of the causative pathogens would aid in the timely initiation and management of ARIs [11, 24].

18 respiratory virus subtypes including coronavirus and human metapneumovirus were included. Of 23,306 swab specimens received, a total of 23,000 pathogens including co-infections were detected by multiplex PCR method. 5 main respiratory pathogens detected in this study was enterovirus/ rhinovirus (29.7%), followed by RSV (15.9%), influenza A (13.7%), adenovirus (11.5%), and parainfluenza virus (10.1%). Noted PCR testing for respiratory pathogens grow at a higher CAGR compared to rapid influenza diagnostic testing, mainly due to higher awareness among the clinicians on the advantage of PCR testing.

Both enterovirus and rhinovirus are within the *Picornaviridae* family. Due to their common genomic features and their high sequence homology, both viruses unfortunately cannot be differentiated by this methodology. Data showed that enterovirus/ rhinovirus was the most predominant pathogens in paediatric ARIs. This is consistent with Yew SM et al. study that showed enterovirus/rhinovirus as the most common virus in the paediatric population, especially among infants (less than 12 months old) [25]. Our study showed that enterovirus/ rhinovirus was detected in 42.1% of the children less than one year old. This finding is consistent with other studies where enterovirus/ rhinovirus are two to three times more frequently detected in infants than older children [26].

This study showed that as the age of the paediatric patients increased, the positive detection rate decreased proportionately from 78% in the age group less than one year old to 58% in the older children group, age 13-17 years old. Studies showed that young children are prone to respiratory infections as their natural passive immunity from the mother diminishes within months after birth and immunological protection will eventually be acquired through infection or from immunisation [27].

Multiple infections were observed in 18.9% of the samples with at least two respiratory pathogens. The significance of co-infections in this study could not be determined as we did not have information on its clinical severity. Some studies have suggested that multiple infections were associated with lengthened duration of hospitalisation, intensive care unit admission, and prolonged mechanical ventilation support and death [28], whereas other studies do not show any differences in severity between single and multiple respiratory infections [29].

The data obtained showed shown that respiratory pathogens were detected year-round. RSV demonstrated the most pronounced seasonality, with peak infection occurring from during July to September. This is supported by a few studies in the region [30-33]. Thongpan et al. in a recent study discovered that RSV infections happened predominantly in the second half of the year between July to November, likely due to the rainy season [30]. Singapore observed similar seasonality patterns with a slight increase in RSV infections between the months of June and August annually [31]. A high proportion of RSV-positive children (25.8%) were observed in infants less than one year old. RSV infected children below 2 years old had a high hospitalisation rate (84.5%). It is also the leading cause of hospitalisation in children less than 2 years old worldwide [32], causing severe pneumonia and bronchiolitis and is associated with a higher risk of developing asthma and recurrent wheezing later in life [33]. Influenza infection from our study, however, was present year-round with no annual seasonality observed. Influenza displays a distinct seasonal pattern in temperate countries, with annual epidemic occurring in the winter. This seasonal pattern however is less commonly reported in tropical or subtropical areas showing year-round circulation with unpredictable variation in intensity [34-35]. Influenza related hospitalisation rate was found to be more prevalent in children less than 6 years old in our study. A similar pattern was also observed where the age-specific hospitalisation rate was higher in younger children less than 5 years old [36].

Given the high RSV disease burden in our younger population, the development of an effective RSV vaccine is of priority. Currently, there is no licensed vaccine for RSV. Several clinical trials are in various stages of development to assess the safety and effectiveness of different RSV vaccine candidates [37]. Understanding the RSV epidemiological patterns helps determine the optimal timing for vaccination. Surveillance of RSV has been integrated into the existing influenza surveillance system in several high-income countries. However, most low- and middle-income countries where the RSV burden is likely to be the greatest has limited or no national data [38]. To our knowledge, this is one of the largest sample sizes of laboratory confirmed influenza and RSV cases in Malaysia

The US Advisory Committee on Immunisation Practices (ACIP) recommends annual influenza vaccination starting from a minimum age of 6 months to reduce influenza-associated morbidity and mortality [39]. Various studies including clinical trials and observational studies have consistently shown that influenza vaccines are safe [40]. However, the complex seasonal patterns of influenza activity in this region have complicated the determination of an optimal timing for influenza vaccination. Our study shows a high rate of influenza infection hospitalisation in children below 6 years of age, with a peak in the 3-6 years old age group. In view of this finding, targeting influenza vaccination in children below 6 years old should be considered.

There are two main limitations in this study. As the data collected was retrospectively and originated from our network of laboratories, the severity and associated comorbidities of the patients could not be assessed. We were also unable to determine the clinical significance of respiratory co-infections. Despite these limitations, the strength of this study is the large sample size providing an insight into the epidemiology and the seasonality of the respiratory pathogens amongst Malaysian paediatric population. Knowledge of the respiratory pathogens epidemiological dynamics will facilitate the identification of a target age and an ideal window for vaccination.

## Data Availability

Data cannot be shared publicly. Data are available from the principle investigator for researchers who meet the criteria for access to confidential data.

## Author’s Contribution

YLL: Conceptualisation, data extraction, formal analysis, visualisation, writing – review & editing

SYW: Conceptualisation, formal analysis, bioinformatic analysis, visualisation, writing – original draft

EKHL: Conceptualisation, visualisation, writing – review & editing

MHM: Conceptualisation, writing – review & editing

## References

[1] Williams BG, Gouws E, Boschi-Pinto C, Bryce J, Dye C. Estimates of worldwide distribution of child deaths from acute respiratory infections. Lancet Infect Dis 2002; 2:25–32.

[2] Assane D, Makhtar C, Abdoulaye D, Amary F, Djibril B, Amadou D, et al. Viral and bacterial etiologies of acute respiratory infections among children under 5 years in Senegal. Microbiol Insights 2018; 11:1–5.

[3] van der Zalm MM, van Ewijk BE, Wilbrink B, Uiterwaal CS, Wolfs TF, van der Ent CK. Respiratory pathogens in children with and without respiratory symptoms. J Pediatr 2009; 154:396–400.

[4] Bicer S, Giray T, Çöl D, Erdağ GÇ, Vitrinel A, Gürol Y, et al. Virological and clinical characterizations of respiratory infections in hospitalized children. Ital J Pediatr 2013; 39:22.

[5] Gülen F, Yildiz B, Çiçek C, Demir E, Tanaç R. Ten-year retrospective evaluation of the seasonal distribution of agent viruses in childhood respiratory tract infections. Turk Pediatri Ars 2014; 49:42–6.

[6] Dong W, Chen Q, Hu Y, He D, Liu J, Yan H, et al. Epidemiological and clinical characteristics of respiratory viral infections in children in Shanghai, China. Arch Virol 2016; 161:1907–13.

[7] Fan J, Henrickson KJ, Savatski LL. Rapid simultaneous diagnosis of infections with respiratory syncytial viruses A and B, influenza viruses A and B, and human parainfluenza virus types 1, 2, and 3 by multiplex quantitative reverse transcription-polymerase chain reaction-enzyme hybridization assay (Hexaplex). Clin Infect Dis 1998; 26:1397–402.

[8] Bellau-Pujol S, Vabret A, Legrand L, Dina J, Gouarin S, Petitjean-Lecherbonnier J, et al. Development of three multiplex RT-PCR assays for the detection of 12 respiratory RNA viruses. J Virol Methods 2005; 126:53–63.

[9] Sloots TP, Whiley DM, Lambert SB, Nissen MD. Emerging respiratory agents: New viruses for old diseases? J Clin Virol 2008; 42:233–43.

[10] Loeffelholz M, Chonmaitree T. Advances in diagnosis of respiratory virus infections. Int J Microbiol 2010; 2010:126049.

[11] Mahony JB. Detection of respiratory viruses by molecular methods. Clin Microbiol Rev 2008; 21:716–47.

[12] Monto AS. Epidemiology of viral respiratory infections. Am J Med 2002; 112:4–12.

[13] Abubakar II, Tillmann T, Banerjee A. Global, regional, and national age-sex specific all-cause and cause-specific mortality for 240 causes of death, 1990-2013: A systematic analysis for the Global Burden of Disease Study 2013. Lancet 2015; 385:117–71.

[14] Piedimonte G, Perez MK. Respiratory syncytial virus infection and bronchiolitis. Pediatr Rev 2014; 35:519–30.

[15] Mancinelli L, Onori M, Concato C, Sorge R, Chiavelli S, Coltella L, et al. Clinical features of children hospitalized with influenza A and B infections during the 2012– 2013 influenza season in Italy. BMC Infect Dis. 2015; 16:1–8.

[16] Hong KW, Cheong HJ, Song JY, Noh JY, Yang TU, Kim WJ. Clinical manifestations of influenza A and B in children and adults at a tertiary hospital in Korea during the 2011–2012 season. Jpn J Infect Dis 2015; 68:20–6.

[17] Bloom-Feshbach K, Alonso WJ, Charu V, Tamerius J, Simonsen L, Miller MA, et al. Latitudinal variations in seasonal activity of influenza and respiratory syncytial virus (RSV): A global comparative review. PLoS One 2013; 14:e54445.

[18] Viboud C, Alonso WJ, Simonsen L. Influenza in tropical regions. PLoS Med 2006; 3:e89.

[19] Park AW, Glass K. Dynamic patterns of avian and human influenza in east and southeast Asia. Lancet Infect Dis 2007; 7:543–8.

[20] Moura FE, Perdigão AC, Siqueira MM. Seasonality of influenza in the tropics: a distinct pattern in northeastern Brazil. Am J Trop Med Hyg 2009; 81:180–3.

[21] Ministry of Health Malaysia Health Fact 2019 https://www.moh.gov.my/moh/resources/Penerbitan/Penerbitan%20Utama/HEALTH%20FACTS/Health%20Facts%202019_Booklet.pdf

[22] Khor CS, Sam IC, Hooi PS, Quek KF, Chan YF. Epidemiology and seasonality of respiratory viral infections in hospitalized children in Kuala Lumpur, Malaysia: a retrospective study of 27 years. BMC Pediatr 2012; 12:1–9.

[23] Saat Z, Abdul Rashid TR, Yusof MA, Kassim FM, Thayan R, Lau SK, et al. Seasonal influenza virus strains circulating in Malaysia from 2005 to 2009. Southeast Asian J Trop Med Public Health 2010; 41:1368–73.

[24] Das S, Dunbar S, Tang YW. Laboratory diagnosis of respiratory tract infections in children – The state of the art. Front Microbiol 2018; 9:2478.

[25] Yew SM, Tan KL, Yeo SK, Ng KP, Kuan CS. Molecular epidemiology of respiratory viruses among Malaysian young children with a confirmed respiratory infection during 2014–2015. J Thorac Dis 2019; 11:4626–33.

[26] Anders KL, Nguyen HL, Nguyen NM, Van Thuy NT, Van NT, Hieu NT, et al. Epidemiology and virology of acute respiratory infections during the first year of life: A birth cohort study in Vietnam. Pediatr Infect Dis 2015; 34:361–70.

[27] Dagne H, Andualem Z, Dagnew B, Taddese AA. Acute respiratory infection and its associated factors among children under-five years attending pediatrics ward at University of Gondar Comprehensive Specialized Hospital, Northwest Ethiopia: Institution-based cross-sectional study. BMC Pediatr 2020; 20:1–7.

[28] Chauhan JC, Slamon NB. The impact of multiple viral respiratory infections on outcomes for critically ill children. Pediatr Crit Care Med 2017; 18:e333–8.

[29] Lim FJ, De Klerk N, Blyth CC, Fathima P, Moore HC. Systematic review and meta-analysis of respiratory viral coinfections in children. Respirology 2016; 21:648–55.

[30] Thongpan I, Vongpunsawad S, Poovorawan Y. Respiratory syncytial virus infection trend is associated with meteorological factors. Sci Rep 2020; 10:1–7.

[31] Lee MW, Goh AE. Mortality in children hospitalised with respiratory syncytial virus infection in Singapore. Singapore Med J 2020.

[32] Lozano R, Naghavi M, Foreman K, Lim S, Shibuya K, Aboyans V, et al. Global and regional mortality from 235 causes of death for 20 age groups in 1990 and 2010: A systematic analysis for the Global Burden of Disease Study 2010. Lancet 2012; 380:2095–128.

[33] Homaira N, Briggs N, Oei JL, Hilder L, Bajuk B, Jaffe A, et al. Association of age at first severe respiratory syncytial virus disease with subsequent risk of severe asthma: a population-based cohort study. J Infect Dis 2019; 220:550–6.

[34] Young BE, Chen M. Influenza in temperate and tropical Asia: A review of epidemiology and vaccinology. Hum Vaccin Immunother 2020; 16:1659–67.

[35] Yuan H, Kramer SC, Lau EH, Cowling BJ, Yang W. Modeling influenza seasonality in the tropics and subtropics. PLoS Comput Biol 2021; 17:e1009050.

[36] Wang X, Li Y, O’Brien KL, Madhi SA, Widdowson MA, Byass P, Omer SB, Abbas Q, Ali A, Amu A, Azziz-Baumgartner E. Global burden of respiratory infections associated with seasonal influenza in children under 5 years in 2018: a systematic review and modelling study. Lancet Glob Health 2020; 8:e497–510.

[37] Soto JA, Stephens LM, Waldstein KA, Canedo-Marroquín G, Varga SM, Kalergis AM. Current insights in the development of efficacious vaccines against RSV. Front Immunol 2020; 11:1507.

[38] Broor S, Campbell H, Hirve S, Hague S, Jackson S, Moen A, et al. Leveraging the Global Influenza Surveillance and Response System for global respiratory syncytial virus surveillance – Opportunities and challenges. Influenza Other Respir Viruses 2020; 14:622–9.

[39] Fiore AE, Uyeki TM, Broder K, Finelli L, Euler GL, Singleton JA, et al. Prevention and control of influenza with vaccines: recommendations of the Advisory Committee on Immunization Practices (ACIP), 2010. MMWR Recomm Rep 2010; 59:1–62.

[40] Sullivan SG, Price OH, Regan AK. Burden, effectiveness and safety of influenza vaccines in elderly, paediatric and pregnant populations. Ther Adv Vaccines Immunother 2019; 7:1–16.

